# Behavioural readiness, not demographics, predicts wearable adoption and digital medicine integration in a diverse multinational population: a cross-sectional study of 3,004 adults in Qatar

**DOI:** 10.64898/2026.07.17.26358328

**Authors:** Hadeel Zaghloul, Batoul Arabi, Minatullah Al-Ani, Aya Abdullah, Reel El-Masri, Ola AboMuslim, Fares Al-Ahdab, Mohamed Rayyan Mohamed Rizwan, Zahir Tag, Shaza Zaghlool, Thurayya Arayssi

## Abstract

Whether diverse populations outside Western settings are behaviourally ready to integrate wearable-derived data into clinical care remains poorly understood. This study examines sociotechnical determinants of wearable adoption and digital health data-sharing readiness in a large, highly diverse multinational population in Qatar, a rapidly digitising health ecosystem with advanced eHealth infrastructure.

We conducted a cross-sectional community-based survey of 3,004 adults across Qatar, assessing wearable device use, behavioural engagement, and willingness to integrate wearable-generated data into healthcare workflows. Multivariable logistic regression identified independent predictors of wearable adoption.

Wearable device use prevalence was 34.1%. Behavioural factors were the strongest independent predictors of adoption: daily exercisers had more than four times the odds of wearable use compared with rarely active participants, and willingness to share data with healthcare providers was independently associated with adoption after full adjustment. Notably, education level was not independently associated with wearable use, suggesting that behavioural readiness outweighs traditional socioeconomic indicators as a determinant of digital health engagement. Older age (≥56 years) and African ethnicity were associated with lower adoption odds, highlighting persistent digital inequities.

These findings challenge the assumption that digital health equity is primarily an education or access problem, repositioning it as a behavioural engagement challenge. Health systems scaling remote monitoring programmes should prioritise identifying behaviourally engaged subpopulations rather than relying solely on demographic targeting. Targeted digital engagement strategies addressing older adults and underrepresented ethnic groups are essential for equitable implementation of digital medicine.

**Author Summary:** Wearable health devices - such as fitness trackers and smartwatches - are increasingly being considered not just as consumer gadgets but as tools that could help doctors monitor patients continuously between clinic visits. For this to work at a population level, we need to understand who is actually using these devices and why. Most research on this question has come from Western countries, leaving large gaps in our understanding of more diverse, rapidly developing health systems.

We surveyed 3,004 adults living in Qatar, one of the most culturally and linguistically diverse countries in the world, and found that about one in three people already uses a wearable health device. More importantly, we found that the strongest predictors of who uses these devices were not education level or socioeconomic status, as is commonly assumed, but rather behavioural factors: how often someone exercises, and whether they are willing to share their device data with a doctor. Daily exercisers were more than four times as likely to use a wearable compared with rarely active individuals, and education level made no difference once we accounted for these behavioural factors.

These findings suggest that efforts to expand the use of digital health tools should focus on identifying and engaging people who are already motivated about their health, rather than assuming that technology training alone will drive adoption. At the same time, older adults and African participants used wearables less often, a reminder that targeted and culturally sensitive approaches remain essential for health equity.

## Introduction

Digital medicine is reshaping healthcare delivery by shifting clinical practice from episodic encounters toward continuous, data-driven models supported by wearable biosensors, remote monitoring, and patient-generated health data streams.(1, 2) Advances in consumer wearables have accelerated this transition by enabling scalable, real-time measurement of physiological and behavioral signals, positioning these devices as potential gateways to precision prevention and personalized care.(3, 4) Despite rapid technological progress, a critical barrier to implementation remains poorly understood: whether diverse populations are behaviorally ready and supported by healthcare systems to integrate wearable-derived data into routine care.(5) Existing research has largely focused on adoption intentions within homogeneous Western populations, leaving a substantial gap in understanding how digital medicine readiness evolves in diverse, rapidly developing healthcare environments outside the northern hemisphere.(6, 7)

Digital health transformation is unfolding unevenly across global regions, creating a need for real-world evidence from populations in evolving healthcare systems.(2, 8) Qatar represents a uniquely informative digital health testbed: a high-income nation with rapid adoption of eHealth infrastructure, a multicultural population with diverse linguistic and socioeconomic backgrounds, and a growing emphasis on precision prevention and remote monitoring.(9, 10) This convergence of advanced digital infrastructure and population heterogeneity offers an opportunity to examine how wearable technologies move beyond consumer novelty toward integration within healthcare delivery.(1, 4) Understanding wearable adoption in such an environment provides insight not only into regional implementation challenges but also into broader questions of scalability, equity, and digital readiness across globally diverse health systems.(3, 11)

Two important gaps shape the motivation for this study. First, prior research on wearable adoption has been predominantly conducted in Western, higher-income, and demographically homogeneous populations, leaving the Middle East and North Africa (MENA) region-where digital health investment is accelerating rapidly-almost entirely uncharacterised at population scale. Second, existing studies have largely treated adoption as a consumer behaviour outcome driven by demographics and socioeconomic status, rather than examining it as a marker of readiness for clinical integration. Rather than examining wearable devices solely as consumer technologies, this study conceptualises wearable adoption as an indicator of digital medicine readiness within a complex, multinational health ecosystem. Critically, we hypothesised that behavioural engagement markers (exercise frequency, data-sharing willingness, and intent to use provider-linked applications) would prove more predictive of adoption than traditional demographic and socioeconomic factors such as education level. If confirmed, this finding would have direct implications for how health systems should design digital health implementation strategies.(1, 4) We aimed to characterize the sociodemographic, health-related, and behavioral determinants that influence wearable health device use and willingness to integrate wearable-derived data into healthcare interactions. By examining associations between wearable adoption and behavioral indicators such as willingness to share data and intent to use provider-linked applications, this study explores factors related to readiness to integrate wearable-derived data into healthcare interactions.(12) Through this approach, the study moves beyond descriptive prevalence to evaluate how population-level factors may shape the transition from lifestyle tracking to clinically integrated digital health infrastructure.(13)

## Results

### Study population characteristics

The study included 3,004 participants (Table 1), of whom 43.0% were male and 57.0% were female. Approximately two-thirds were aged 18-35 years, and a majority identified as Arab. Over three-quarters had at least a college-level education, and about two thirds were never married.

**Table 1.** Characteristics of study participants.

| Characteristics | Total<br>N (%) |
| --- | --- |
| **I. Sociodemographic characteristics** |  |
| **Gender** |  |
| Male | 1,270 (43.0%) |
| Female | 1,686 (57.0%) |
| **Age (years)** |  |
| 18–25 | 961 (32.6%) |
| 26–35 | 964 (32.7%) |
| 36–45 | 641 (21.7%) |
| 46–55 | 275 (9.3%) |
| ≥56 | 108 (3.7%) |
| **Ethnicity** |  |
| Arab | 1,213 (41.1%) |
| African | 420 (14.2%) |
| East Asian | 262 (8.9%) |
| Hispanic | 46 (1.6%) |
| Other | 181 (6.1%) |
| South Asian | 739 (25.0%) |
| White-Caucasian | 93 (3.1%) |
| **Education level** |  |
| High school or less | 676 (22.8%) |
| College or higher | 2,284 (77.2%) |
| **Ever married** |  |
| No | 1,745 (58.9%) |
| Yes | 1,219 (41.1%) |
| **II. Health status and conditions** |  |
| **Chronic conditions** |  |
| No | 2,452 (83.1%) |
| Yes | 497 (16.9%) |
| **Self-rated health** |  |
| Poor-Fair | 577 (19.5%) |
| Good | 1,998 (67.5%) |
| Excellent | 384 (13.0%) |
| **Number of annual doctor visits** |  |
| Never | 372 (12.6%) |
| 1–5 | 2,233 (75.3%) |
| ≥6 | 359 (12.1%) |
| **Perceived weight** |  |
| Underweight | 410 (13.8%) |
| Normal | 1,291 (43.5%) |
| Overweight | 1,264 (42.6%) |
| **III. Behavioral factors** |  |
| **Exercise frequency** |  |
| Rarely | 860 (29.2%) |
| Once a week | 634 (21.5%) |
| Several times a week | 1,152 (39.1%) |
| Everyday | 299 (10.2%) |
| **Exercise enjoyment** |  |
| Not at all | 279 (9.5%) |
| A little | 709 (24.1%) |
| A moderate amount | 1,107 (37.6%) |
| A lot | 849 (28.8%) |
| **Willingness to share smartwatch health data with health care provider** |  |
| No | 915 (31.4%) |
| Yes | 1,997 (68.6%) |
| **Intent to use health care provider app on smartwatch** |  |
| Unlikely | 305 (10.4%) |

| Characteristics | Total |
| --- | --- |
|  | N (%) |
| Neutral | 631 (21.5%) |
| Likely | 1,994 (68.1%) |
| **Total** | **3,004 (100.0%)** |

Missing values were excluded from the analysis.

Most reported having no chronic conditions, rated their health as good, and had between 1-5 doctor visits annually. Additionally, 42.6% reported being overweight.

About half exercised several times per week or more, and 66.4% reported enjoying exercise. Overall, 68.6% were willing to share smartwatch health data with a health care provider, and a similar proportion indicated they were likely to use a provider-specific smartwatch application. Most also reported performing at least two health-related tasks using a computer, smartphone, or other electronic device (Figure 2).

**Figure 1.**
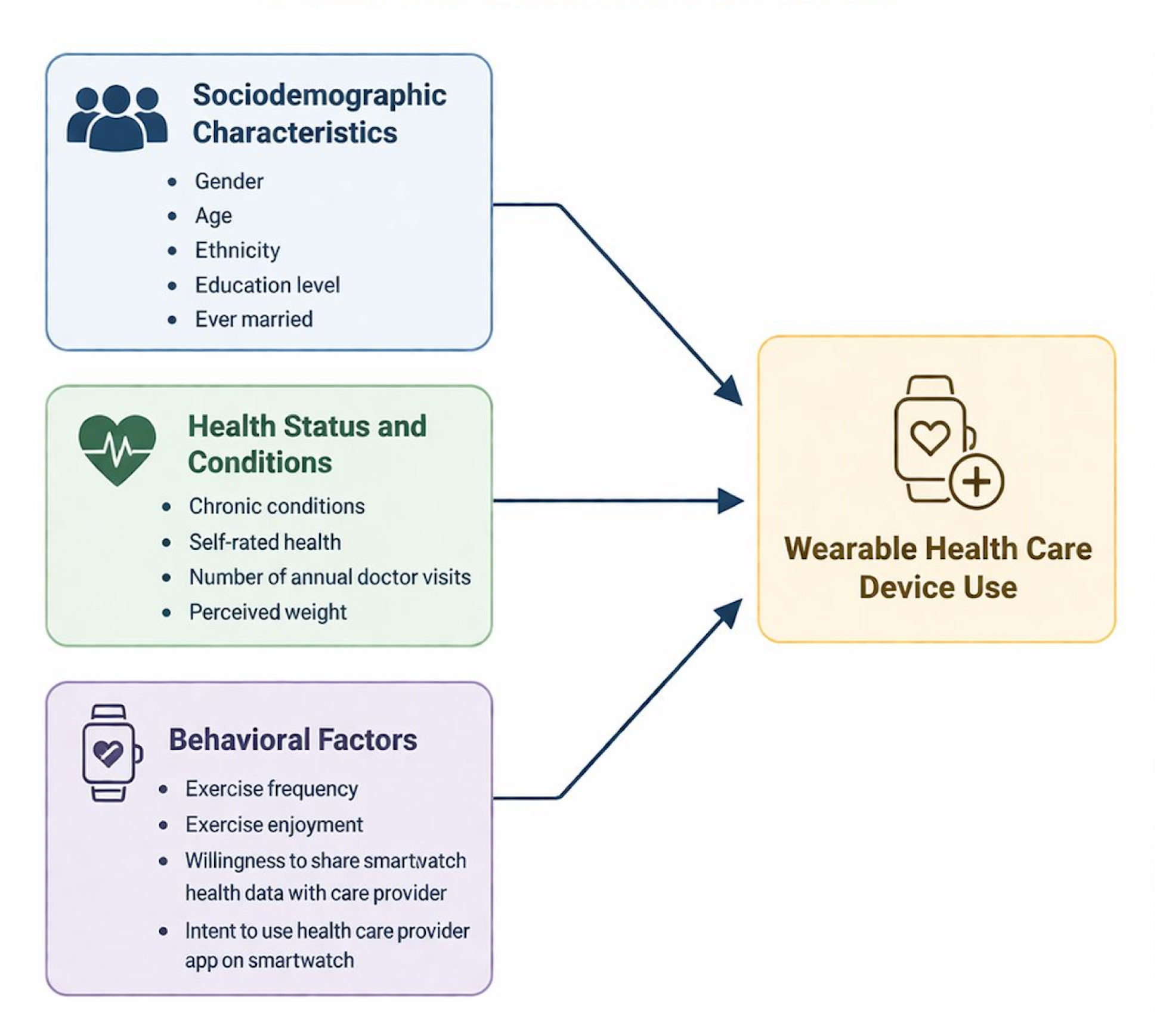
Analytical framework for examining predictors of wearable health care device use.

**Figure 2.**
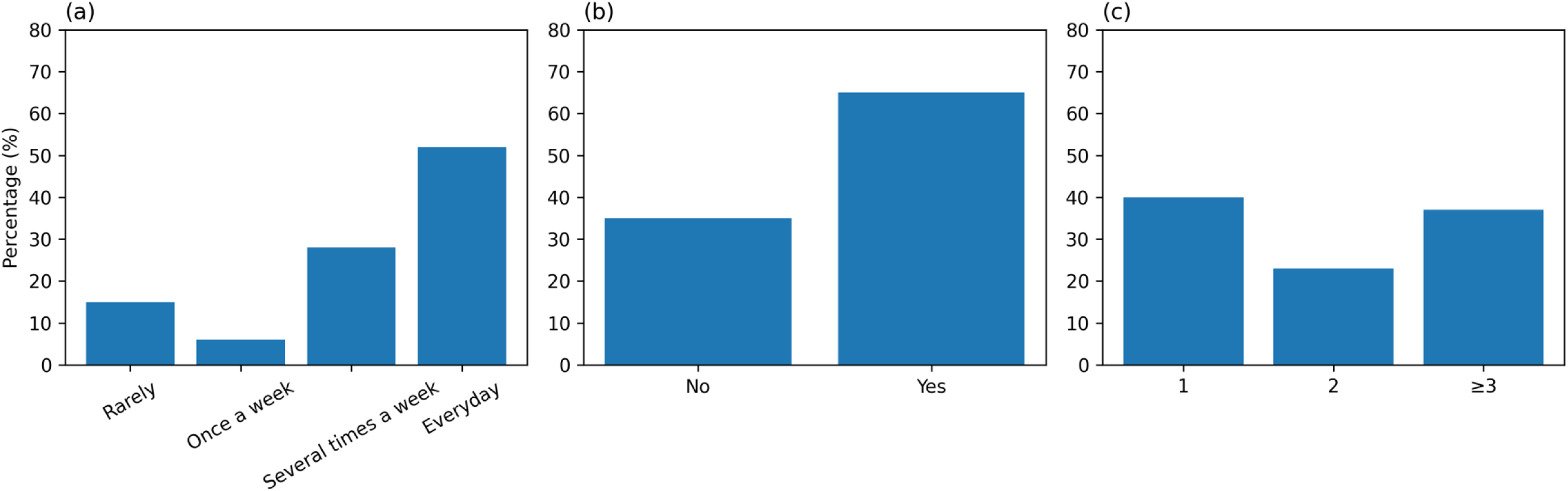
Distribution of participant responses: (a) frequency of wearable health care device use, (b) perceived positive health and lifestyle changes since using a wearable health care device, and (c) number of health-related tasks performed on a computer, smartphone, or other electronic device*. *Health-related tasks include any combination of the following: looking for health information; purchasing medicines or vitamins online; communicating with a provider via email or the internet; tracking health care changes; viewing medical test results; and making appointments with a provider.

### Prevalence of wearable health care device use

The prevalence of wearable health care device use among participants was 34.1% (n= 1024) (95% CI: 32.4-35.9%) (Table 2). Among wearable health care device owners, more than half reported using their device daily (Figure 2), and just over 60% reported positive health and lifestyle changes following use.

**Table 2.** Associations of characteristics with wearable health care device use.

| Characteristics | Total | Wearable health care device use |  |
| --- | --- | --- | --- |
|  | N (%)<sup>a</sup> | N (%)<sup>a</sup> | p-value |
| I. Sociodemographic characteristics |  |  |  |
| Gender |  |  |  |
| Male | 1,264 (42.9%) | 438 (34.7%) | 0.641 |
| Female | 1,682 (57.1%) | 569 (33.8%) |  |
| Age (years) |  |  |  |
| 18–25 | 960 (32.7%) | 338 (35.2%) | 0.003 |
| 26–35 | 958 (32.6%) | 338 (35.3%) |  |
| 36–45 | 640 (21.8%) | 223 (34.8%) |  |
| 46–55 | 274 (9.3%) | 86 (31.4%) |  |
| ≥56 | 107 (3.6%) | 18 (16.8%) |  |
| Ethnicity |  |  |  |
| Arab | 1,209 (41.1%) | 411 (34.0%) | 0.004 |
| African | 419 (14.2%) | 112 (26.7%) |  |
| East Asian | 261 (8.9%) | 94 (36.0%) |  |
| Hispanic | 45 (1.5%) | 19 (42.2%) |  |
| Other | 181 (6.2%) | 71 (39.2%) |  |
| South Asian | 737 (25.0%) | 255 (34.6%) |  |
| White-Caucasian | 91 (3.1%) | 41 (45.1%) |  |
| Education level |  |  |  |
| High school or less | 671 (22.8%) | 213 (31.7%) | 0.135 |
| College or higher | 2,278 (77.2%) | 794 (34.9%) |  |
| Ever married |  |  |  |
| No | 1,738 (58.9%) | 610 (35.1%) | 0.187 |
| Yes | 1,215 (41.1%) | 398 (32.8%) |  |
| II. Health status and conditions |  |  |  |
| Chronic conditions |  |  |  |
| No | 2,444 (83.2%) | 843 (34.5%) | 0.368 |
| Yes | 494 (16.8%) | 160 (32.4%) |  |
| Self-rated health |  |  |  |
| Poor-Fair | 577 (19.6%) | 169 (29.3%) | 0.025 |
| Good | 1,989 (67.5%) | 701 (35.2%) |  |
| Excellent | 382 (13.0%) | 135 (35.3%) |  |
| Number of annual doctor visits |  |  |  |
| Never | 372 (12.6%) | 124 (33.3%) | 0.944 |
| 1–5 | 2,226 (75.4%) | 761 (34.2%) |  |
| ≥6 | 355 (12.0%) | 122 (34.4%) |  |
| Perceived weight |  |  |  |
| Underweight | 407 (13.8%) | 118 (29.0%) | 0.049 |
| Normal | 1,284 (43.5%) | 457 (35.6%) |  |
| Overweight | 1,263 (42.8%) | 433 (34.3%) |  |
| III. Behavioral factors |  |  |  |
| Exercise frequency |  |  |  |
| Rarely | 858 (29.3%) | 157 (18.3%) | <0.001 |
| Once a week | 628 (21.4%) | 211 (33.6%) |  |
| Several times a week | 1,150 (39.2%) | 484 (42.1%) |  |
| Everyday | 296 (10.1%) | 149 (50.3%) |  |
| Exercise enjoyment |  |  |  |
| Not at all | 279 (9.5%) | 57 (20.4%) | <0.001 |
| A little | 707 (24.1%) | 197 (27.9%) |  |
| A moderate amount | 1,102 (37.6%) | 398 (36.1%) |  |
| A lot | 843 (28.8%) | 347 (41.2%) |  |
| Willingness to share smartwatch health data with health care provider |  |  |  |
| No | 909 (31.4%) | 217 (23.9%) | <0.001 |
| Yes | 1,990 (68.6%) | 782 (39.3%) |  |
| Intent to use health care provider app on smartwatch |  |  |  |
| Unlikely | 305 (10.5%) | 60 (19.7%) | <0.001 |
|  | N (%) | N (%) | p-value |
| Neutral | 627 (21.5%) | 193 (30.8%) |  |
| Likely | 1,985 (68.0%) | 746 (37.6%) |  |
| **Total** | **2,957 (100.0%)** | **1,009 (34.1%)** | **NA** |

Abbreviations: NA, not applicable.
\*Missing values were excluded from the analysis.

Prevalence varied by age, peaking in younger age groups and declining with older age (Table 2 and Figure 3). Among ethnic groups, wearable use was most common among Arab participants, who constituted the majority of the sample, and was also high among White/Caucasian and Hispanic participants, while the lowest prevalence was observed among African participants. Prevalence was also higher among those who rated their health more favorably, had normal weight or were overweight, exercised more frequently, reported greater enjoyment of exercise, were willing to share wearables device health data with a health care provider, and were more likely to use a provider-specific smartwatch application.

**Figure 3.**
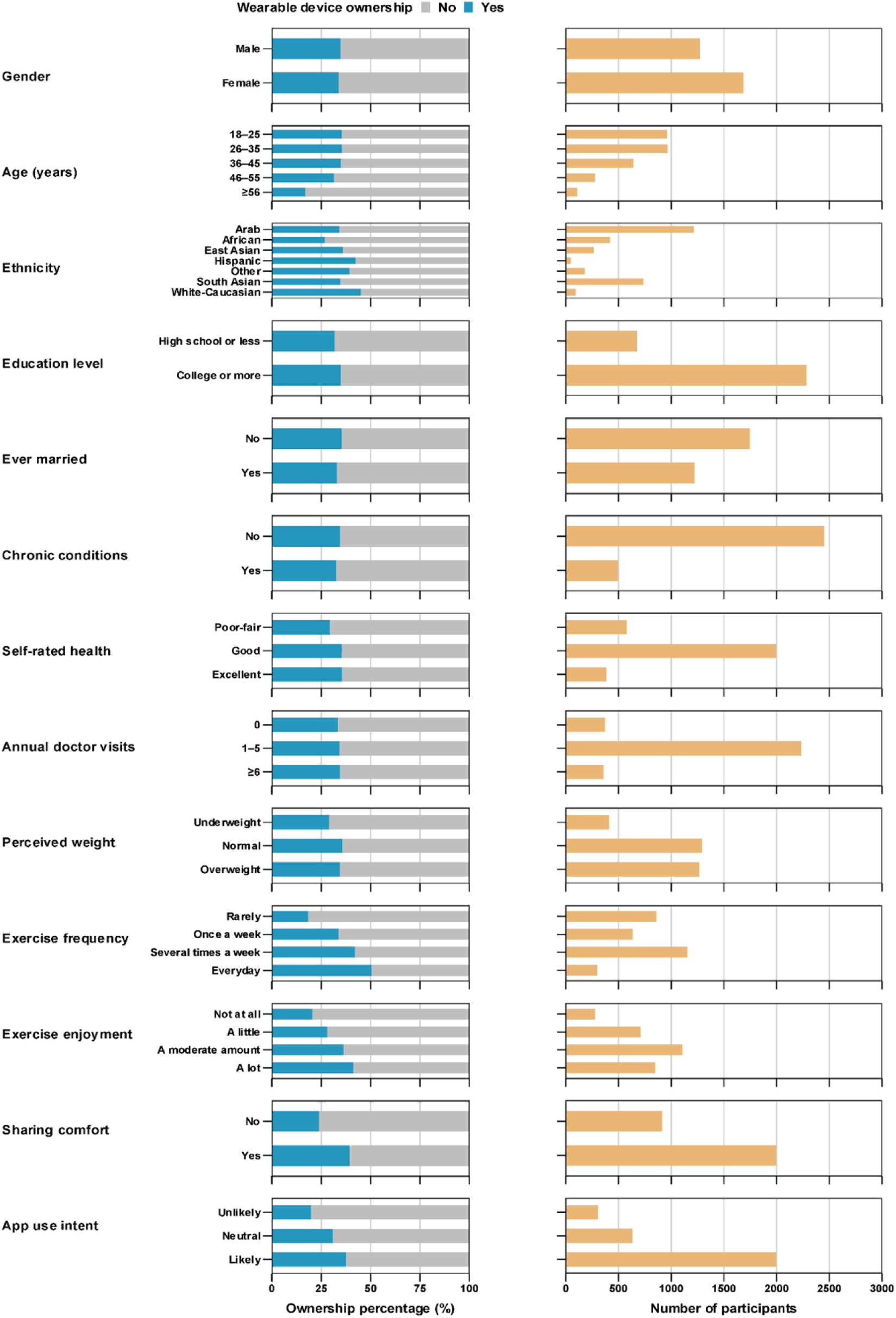
Wearable health care device use across sociodemographic, health, and behavioral factors. In the figure, “annual doctor visits” refers to the number of annual doctor visits; “sharing comfort” refers to willingness to share smartwatch health data with a health care provider; and “app use intent” refers to intent to use a health care provider app on a smartwatch.

### Factors associated with wearable health care device use

Results of the multivariable logistic regression analysis are presented in Table 3 and illustrated in Figure 4. Variance inflation factor (VIF) was <5 in the model, indicating no multicollinearity among predictor variables.

**Table 3.** Factors associated with wearable health care device use.

| Characteristics | Univariable regression analysis |  |  | Multivariable regression analysis |  |
| --- | --- | --- | --- | --- | --- |
|  | OR (95% CI) | p-value | LR test p-value* | aOR (95% CI) | p-value† |
| **I. Sociodemographic** |  |  |  |  |  |
| **Gender** |  |  |  |  |  |
| Male | 1.00 |  | 0.641 | -- | -- |
| Female | 0.96 (0.83-1.12) | 0.641 |  | -- | -- |
| **Age (years)** |  |  |  |  |  |
| 18-25 | 1.00 |  | 0.001 | 1.00 |  |
| 26-35 | 1.00 (0.83-1.21) | 0.973 |  | 1.14 (0.92-1.42) | 0.230 |
| 36-45 | 0.98 (0.80-1.21) | 0.881 |  | 1.07 (0.82-1.40) | 0.614 |
| 46-55 | 0.84 (0.63-1.12) | 0.240 |  | 0.94 (0.67-1.33) | 0.730 |
| ≥56 | 0.37 (0.22-0.63) | <0.001 |  | 0.44 (0.25-0.78) | 0.005 |
| **Ethnicity** |  |  |  |  |  |
| Arab | 1.00 |  | 0.004 | 1.00 |  |
| African | 0.71 (0.55-0.91) | 0.006 |  | 0.71 (0.54-0.92) | 0.010 |
| East Asian | 1.09 (0.83-1.45) | 0.533 |  | 1.15 (0.84-1.56) | 0.384 |
| Hispanic | 1.42 (0.78-2.59) | 0.256 |  | 1.45 (0.76-2.78) | 0.259 |
| Other | 1.25 (0.91-1.73) | 0.168 |  | 1.20 (0.85-1.70) | 0.290 |
| South Asian | 1.03 (0.85-1.25) | 0.785 |  | 1.07 (0.87-1.33) | 0.508 |
| White-Caucasian | 1.59 (1.04-2.45) | 0.034 |  | 1.77 (1.11-2.82) | 0.016 |
| **Education level** |  |  |  |  |  |
| High school or less | 1.00 |  | 0.134 | 1.00 |  |
| College or higher | 1.15 (0.96-1.38) | 0.135 |  | 0.93 (0.75-1.15) | 0.497 |
| **Ever married** |  |  |  |  |  |
| No | 1.00 |  | 0.186 | 1.00 |  |
| Yes | 0.90 (0.77-1.05) | 0.187 |  | 0.95 (0.77-1.17) | 0.621 |
| **II. Health status and conditions** |  |  |  |  |  |
| **Chronic conditions** |  |  |  |  |  |
| No | 1.00 |  | 0.367 | -- | -- |
| Yes | 0.91 (0.74-1.12) | 0.369 |  | -- | -- |
| **Self-rated health** |  |  |  |  |  |
| Poor-Fair | 1.00 |  | 0.024 | 1.00 |  |
| Good | 1.31 (1.07-1.61) | 0.008 |  | 1.09 (0.88-1.37) | 0.429 |
| Excellent | 1.32 (1.00-1.74) | 0.049 |  | 0.95 (0.69-1.29) | 0.732 |
| **Number of annual doctor visits** |  |  |  |  |  |
| Never | 1.00 |  | 0.944 | -- | -- |
| 1-5 | 1.04 (0.82-1.31) | 0.748 |  | -- | -- |
| ≥6 | 1.05 (0.77-1.42) | 0.769 |  | -- | -- |
| **Perceived weight** |  |  |  |  |  |
| Underweight | 1.00 |  | 0.047 | 1.00 |  |
| Normal | 1.35 (1.06-1.73) | 0.015 |  | 1.26 (0.97-1.63) | 0.087 |
| Overweight | 1.28 (1.00-1.63) | 0.049 |  | 1.52 (1.16-1.98) | 0.002 |
| **III. Behavioral factors** |  |  |  |  |  |
| **Exercise frequency** |  |  |  |  |  |
| Rarely | 1.00 |  | <0.001 | 1.00 |  |
| Once a week | 2.26 (1.78-2.87) | <0.001 |  | 2.18 (1.69-2.82) | <0.001 |
| Several times a week | 3.24 (2.63-4.00) | <0.001 |  | 2.88 (2.27-3.65) | <0.001 |
| Everyday | 4.53 (3.40-6.02) | <0.001 |  | 4.21 (3.05-5.82) | <0.001 |
| **Exercise enjoyment** |  |  |  |  |  |
| Not at all | 1.00 |  | <0.001 | 1.00 |  |
| A little | 1.50 (1.08-2.10) | 0.017 |  | 1.10 (0.76-1.58) | 0.615 |
| A moderate amount | 2.20 (1.61-3.02) | <0.001 |  | 1.22 (0.86-1.75) | 0.267 |
| A lot | 2.72 (1.98-3.76) | <0.001 |  | 1.26 (0.86-1.83) | 0.235 |
| Willingness to share smartwatch health data with health care provider |  |  |  |  |  |
| No | 1.00 |  | <0.001 | 1.00 |  |
| Yes | 2.06 (1.73-2.46) | <0.001 |  | 1.78 (1.46-2.17) | <0.001 |
| Intent to use health care provider app on smartwatch |  |  |  |  |  |
| Unlikely | 1.00 |  | <0.001 | 1.00 |  |
| Neutral | 1.82 (1.31-2.52) | <0.001 |  | 1.48 (1.05-2.10) | 0.027 |
| Likely | 2.46 (1.83-3.31) | <0.001 |  | 1.68 (1.21-2.33) | 0.002 |

Abbreviations: aOR, adjusted odds ratio; CI, confidence interval; LR, likelihood ratio; OR, odds ratio.
\*Covariates with p-value $\leq 0.2$ in the univariable analysis were included in the multivariable analysis.
†Covariates with p-value $< 0.05$ in the multivariable analysis were considered as showing statistically significant evidence for an association with wearable health care device use.

**Figure 4.**
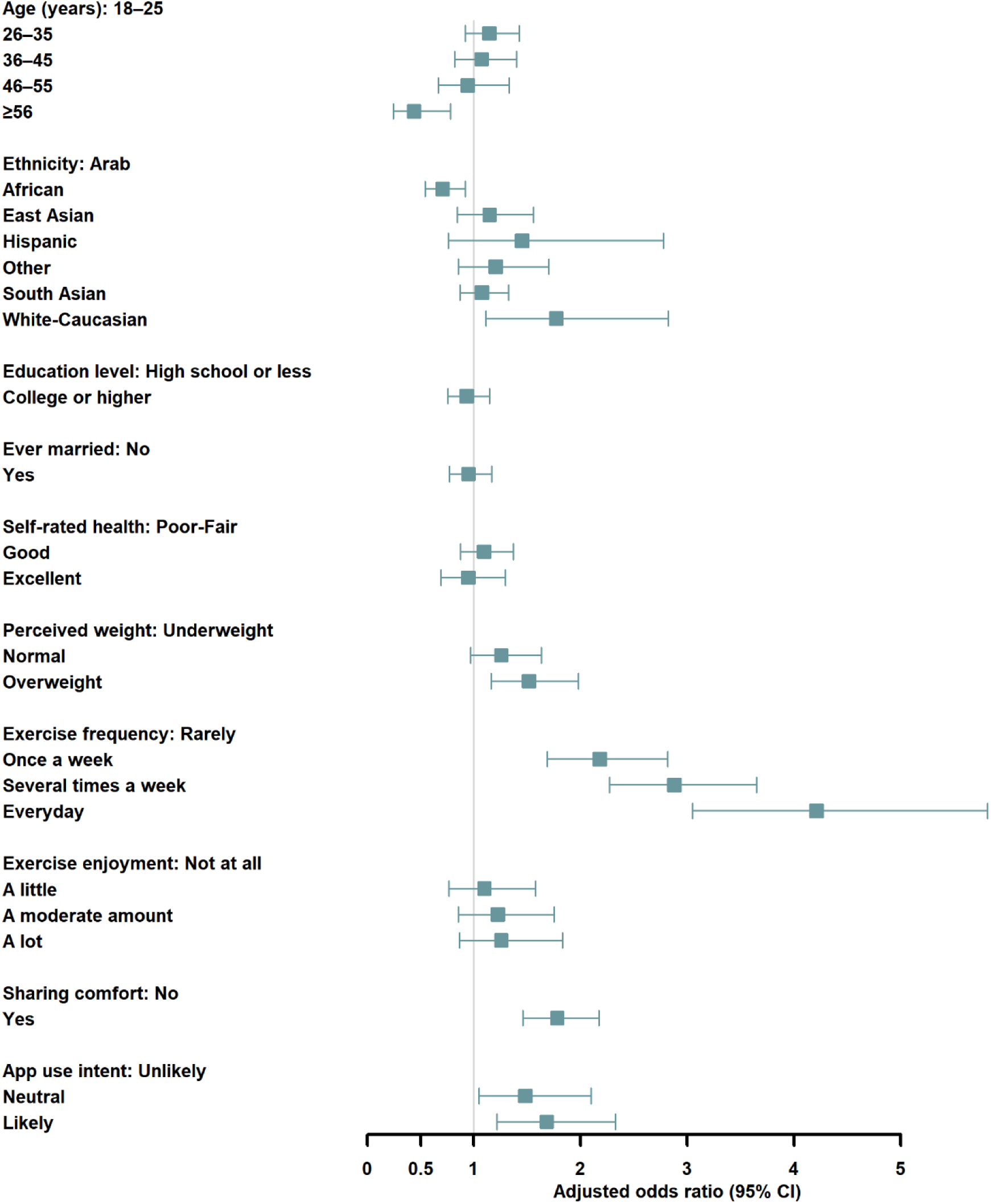
Forest plot presenting multivariable regression analysis results of factors associated with wearable health care device use. Abbreviations: CI, confidence interval. In the figure, “sharing comfort” refers to willingness to share smartwatch health data with a health care provider, and “app use intent” refers to intent to use a health care provider app on a smartwatch.

#### Sociodemographic characteristics

Participants aged 56 years or older had 56% lower odds of wearable health care device use compared with those aged 18-25 years. Africans had the lowest odds compared with Arabs (aOR: 0.71; 95% CI: 0.54-0.92), while White/Caucasian participants had the highest odds (only 3%) (aOR: 1.77; 95% CI: 1.11-2.82). No associations were found for gender, education level, and marital status.

#### Health status and conditions

Participants who were overweight had higher odds of wearable health care device use compared with those who were underweight (aOR: 1.52; 95% CI: 1.16-1.98). No associations were found for chronic conditions, self-rated health, and number of annual doctor visits.

#### Behavioral factors

Compared with those who rarely exercised, participants who exercised once a week (aOR: 2.18; 95% CI: 1.69-2.82), several times a week (aOR: 2.88; 95% CI: 2.27-3.65), or daily (aOR: 4.21; 95% CI: 3.05-5.82) had progressively higher odds of wearable health care device use. Higher odds were also observed among those willing to share smartwatch health data with a health care provider (aOR: 1.78; 95% CI: 1.46-2.17). Additionally, compared with those unlikely to use a provider-specific smartwatch application, participants who were neutral (aOR: 1.48; 95% CI: 1.05-2.10) or likely (aOR: 1.68; 95% CI: 1.21-2.33) had higher odds of wearable health care device use. No association was found for exercise enjoyment.

## Discussion

This study positions wearable health device adoption as a measurable indicator of digital medicine readiness within a diverse, rapidly digitizing population.(2, 14, 15) Our findings suggest that wearable use reflects higher engagement in health-related behaviors and greater readiness to use digital health tools, rather than just ownership of consumer technology, with potential relevance for remote monitoring and prevention strategies.(16, 17) As digital health transitions from episodic care toward continuous sensing and personalized feedback, understanding these sociotechnical determinants becomes essential for equitable implementation across heterogeneous populations.(18, 19)

### Sociodemographic patterns and the digital divide

Consistent with global evidence, wearable adoption declined significantly with increasing age.(20) Older adults demonstrated markedly lower odds of device use, highlighting persistent digital inequities that could widen as healthcare systems increasingly rely on patient-generated data streams. This finding echoes broader concerns about the digital divide, where structural factors such as technological literacy, accessibility, and perceived relevance influence adoption patterns.(21, 22) While younger populations may naturally adopt wearable technologies, older adults, who often carry a higher burden of chronic disease, risk being excluded from the benefits of digital medicine if implementation strategies fail to address usability, education, and cultural barriers.(23)

The absence of education level as an independent predictor in the adjusted model is perhaps the most important and counterintuitive finding of this study. Digital health equity has traditionally been framed as a problem of access and literacy - the assumption being that better-educated individuals adopt digital health tools at higher rates because they can understand and navigate them more effectively. Our data directly challenge this framing. After accounting for behavioural factors, education explained no additional variance in wearable adoption. This repositions the digital divide not primarily as a literacy or access problem, but as a motivational and behavioural engagement challenge. The implication for health systems is substantial: investing in digital literacy programmes alone is unlikely to close adoption gaps if the underlying barriers are attitudinal and behavioural rather than cognitive. Implementation strategies that promote health motivation, establish clear perceived utility for wearables in clinical care, and build patient trust in data sharing may be more effective than those focused on technology training alone. This finding aligns with emerging research indicating that digital engagement behaviours and health motivation are more influential than demographic characteristics in determining adoption trajectories,(24, 25) For healthcare systems planning large-scale digital interventions, these findings underscore the importance of targeting behavioural readiness rather than relying solely on demographic proxies. Critically, this evidence now extends to a MENA population for the first time, providing a regional anchor for digital health implementation frameworks in the Gulf and broader Arab world.

Two other findings in the adjusted model warrant specific discussion. First, African participants had significantly lower odds of wearable adoption compared with Arab participants, even after full adjustment for sociodemographic and behavioural factors (aOR: 0.71; 95% CI: 0.54–0.92). This persistent disparity is not fully explained by the variables measured in this study and points toward unmeasured structural determinants, potentially including device cost, language barriers in device interfaces, differential social norms around health self-monitoring, or lower trust in digital data sharing systems. These possibilities cannot be disentangled with cross-sectional data, and future qualitative and longitudinal research specifically examining wearable adoption barriers among African communities in Qatar is warranted. From an implementation standpoint, this finding signals that universal digital health roll-out strategies are unlikely to be equitable without targeted outreach and culturally adapted engagement approaches for underrepresented ethnic groups. Second, being overweight was independently associated with higher wearable adoption odds compared with being underweight. Rather than reflecting a paradox, this finding is likely explained by health-motivated monitoring: individuals with elevated weight may be more actively engaged in self-monitoring behaviours as part of weight management or chronic disease prevention efforts. This interpretation is consistent with the broader pattern observed in this study, that wearable adoption is driven by proactive health engagement rather than clinical need alone, and reinforces the behavioural readiness framing central to our conceptual framework.

### Digital medicine readiness and behavioral engagement

One of the most consistent findings in this study was the strong association between wearable adoption and behavioral engagement, particularly exercise frequency and willingness to share health data with healthcare providers. This aligns with prior research showing that wearable technologies are most effective when embedded within behavior change ecosystems rather than functioning as passive monitoring tools.(26, 27) Digital behavior change interventions incorporating self-monitoring, feedback, and goal-setting have demonstrated significant improvements in physical activity outcomes across randomized trials, supporting the role of engagement-driven design in wearable effectiveness.(28) Individuals who exercised more frequently or expressed readiness to integrate device-generated data into clinical workflows demonstrated substantially higher adoption, suggesting that wearable use may reflect a broader orientation toward proactive health management. Similar patterns have been reported in digital health adoption studies, where perceived utility, self-efficacy, and engagement behaviors strongly predict sustained use of digital technologies.(29)

Importantly, our results extend this literature by demonstrating these relationships in a multicultural population outside Western contexts.(25, 30) The high proportion of participants willing to share wearable data with providers suggests emerging acceptance of digitally mediated care, reinforcing the notion that behavioral readiness may serve as an early signal for successful integration of remote monitoring systems.(25, 31) From a digital medicine perspective, these findings emphasize that scalable digital health solutions require not only technological capability but also strategies that actively promote user engagement and trust. (32, 33)

### Health system integration and implementation implications

Beyond individual-level predictors, this study suggests an emerging alignment between consumer wearable use and attitudes toward healthcare integration, as demonstrated by the strong associations observed between wearable use and willingness to share health data with providers, as well as intent to use provider-linked smartwatch applications. In particular, participants who reported willingness to share wearable-derived data and those expressing intent to use provider linked applications had significantly higher odds of wearable device use in the multivariable analysis, indicating that these behavioral markers may reflect readiness for integration into clinical workflows. The observed associations between wearable device use and both willingness to share health data with healthcare providers and intent to use provider-linked smartwatch applications suggest that individuals who adopt wearable technologies may be more receptive to models that incorporate these data into clinical care. Evidence from remote patient monitoring studies shows that perceived clinical utility and interoperability strongly influence wearable uptake and sustained engagement.(1, 34) As wearable technologies evolve from lifestyle tools toward clinically relevant sensors, patient perceptions of medical utility may increasingly shape uptake.(35, 36)

By describing associations between wearable adoption and key markers of behavioral readiness, including younger age, higher exercise frequency, willingness to share health data with healthcare providers, and intent to use provider-linked smartwatch applications, all of which were independently associated with wearable use in the multivariable analysis, this study provides empirical context for understanding how digital health tools are adopted in real world settings. Implementation science research emphasizes that successful scaling of digital health innovations depends on identifying digitally engaged subgroups and aligning technology deployment with behavioral readiness and system-level capacity.(11, 37) In rapidly developing healthcare systems such as Qatar’s, where investments in eHealth infrastructure and precision prevention initiatives are accelerating, identifying early adopters may inform targeted remote monitoring pilots and implementation strategies. Similar approaches have been proposed in digital medicine frameworks, where early adopters act as catalysts for broader system integration while digital literacy interventions address adoption gaps across populations.(30) By describing associations between markers of behavioral readiness and wearable adoption, this study provides empirical context for discussions on how digital health tools are taken up in real-world settings.(11) Based on these findings, potential early adopers of digitally integrated healthcare in this population may include younger individuals, those who engage in regular physical activity, and individuals who demonstrate openness to digital health integration through willingness to share data and use provider-linked applications. These groups may represent key entry points for implementing scalable remote monitoring interventions.

### Study context: Digital health environment in Qatar

Qatar provides a unique context for examining wearable adoption within a rapidly evolving digital health ecosystem, characterized by high internet penetration, widespread smartphone use, and a highly multicultural population. (9) (38) This setting enables the evaluation of digital medicine readiness across diverse sociocultural and linguistic groups, offering insights that extend beyond more homogeneous study populations.

Compared with studies conducted in more demographically homogeneous settings, this research captures adoption patterns within a population characterized by high mobility, linguistic diversity, and variable levels of digital literacy. Qatar’s high internet penetration and widespread mobile connectivity provide conditions in which consumer wearable technologies are readily accessible.(39, 40) In a nationally representative sample of U.S. adults, about 30% of adults reported using wearable health care devices.(17) The prevalence observed in this study (34.1%) may reflect high levels of digital access, smartphone penetration exceeding 90%, and ongoing national investment in digital health initiatives.(41)

At the same time, disparities across demographic groups indicate that access to infrastructure alone does not ensure equitable adoption. Prior digital health research in Qatar underscores the importance of culturally adaptive implementation strategies and user-centered design, particularly in multicultural expatriate populations where language, literacy, and sociotechnical factors influence engagement with wearable and remote monitoring technologies.(38)

### Strengths and limitations

A major strength of this study is its large, community-based sample drawn from multiple public settings, allowing for the capture of real-world wearable adoption patterns across a broad demographic spectrum. The multilingual survey design further enhances inclusivity and reflects the sociocultural diversity of the study population. Additionally, the use of multivariable modeling enables identification of independent predictors of wearable adoption, contributing to a nuanced understanding of digital medicine readiness.

Nevertheless, several limitations should be acknowledged. First, the cross-sectional design precludes causal inference. A particular concern is directionality: our data cannot determine whether wearable adoption drives willingness to share data with providers, or whether pre-existing openness to digital health care drives adoption. Longitudinal studies are needed to determine whether behavioural readiness predicts sustained wearable engagement and improved health outcomes over time. Second, wearable use and all predictor variables were self-reported, introducing the possibility of recall and social desirability bias; objectively measured adoption data would strengthen future work. Third, convenience sampling in public venues introduces selection bias. Populations less likely to frequent shopping malls, parks, and cultural centres, including elderly adults, individuals with mobility limitations, and those with very low digital literacy, are likely under-represented, and the findings should be interpreted with this in mind. The very low representation of adults aged 56 and above (3.7% of the sample) limits the precision of estimates for this clinically important group. Fourth, the study describes associations between adoption and behavioural readiness markers; it does not evaluate actual integration of wearable-derived data into clinical workflows or its impact on health outcomes, which remains an important gap for future research. This study was conducted and reported in accordance with the STROBE guidelines for cross-sectional observational research.

### Future directions

Future research should move beyond adoption metrics toward evaluating how wearable-derived data can be meaningfully integrated into healthcare systems to improve clinical outcomes. Longitudinal studies examining digital engagement trajectories, data-sharing behaviors, and health outcomes could provide deeper insight into the mechanisms linking wearable use with precision prevention. Additionally, integrating objective wearable sensor data with electronic health records may help identify digital biomarkers and enhance predictive modeling approaches central to digital medicine. Policymakers and health system leaders should also consider embedding digital literacy and engagement strategies within chronic disease programs to ensure that technological innovation translates into equitable healthcare transformation.

## Conclusion

In a rapidly evolving digital health landscape, wearable adoption reflects more than consumer technology trends; it is an emerging and measurable marker of population-level readiness for digitally integrated care. The central finding of this study, that behavioural engagement predicts adoption more strongly than education or socioeconomic status, challenges a foundational assumption of digital health equity frameworks and has direct implications for how health systems should design implementation strategies. Targeting behaviourally engaged subpopulations, rather than relying on demographic proxies, is likely to yield more equitable and scalable digital health integration. At the same time, the persistent adoption gaps observed among older adults and African participants signal that access-focused and culturally adapted interventions remain necessary alongside behavioural approaches. This study provides, to our knowledge, the first large-scale population-level evidence on digital medicine readiness from the MENA region, offering a reference point for other rapidly digitising health systems in the Gulf, broader Middle East, and comparable global settings. As healthcare increasingly relies on continuous patient-generated data, understanding the human factors that shape engagement will be as critical as the technologies themselves.

## Methods

### Study design and setting

We conducted a cross-sectional community survey to examine patterns of use and determinants of wearable healthcare device adoption among adults in Qatar. Data were collected between January 6, 2025 and July 26 2025 at multiple public venues, including shopping malls, parks, beaches, universities, and cultural centers, to capture a diverse sample of the population.

### Participants and eligibility

Adults aged 18 years or older, residing in Qatar, and able to read and understand English, Arabic, Tamil, Telugu, or Sinhala were eligible. Individuals younger than 18 years or not living in Qatar were excluded. Participation was voluntary, and no incentives were provided. These languages were selected to reflect the major linguistic groups within Qatar’s diverse population, particularly among long-term expatriate communities, to maximize inclusivity and representativeness of the sample.

### Recruitment and sampling strategy

A convenience sampling approach was used. Trained researchers approached individuals in predefined zones of each recruitment site at rotating times of day (morning, afternoon, evening) and across weekdays and weekends to maximize diversity of participants. To maintain flow, one researcher introduced the study and confirmed eligibility, while a second assisted participants in completing the survey. Recruitment was conducted individually in semi-private areas of public spaces to safeguard confidentiality.

### Data collection and survey instrument

Data were collected on institution-issued iPads using Qualtrics survey software. The survey was available in five languages (English, Arabic, Tamil, Telugu, Sinhala) and required approximately 20 minutes to complete. The first screen presented the study information sheet; proceeding to the next page was considered provision of electronic informed consent. Participants were informed they could withdraw at any time before submitting responses. No personal identifiers were collected.

The survey included the following domains:

- Wearable use: current use of a wearable healthcare device (defined in the survey as a fitness tracker or smartwatch with health features), frequency of use, and perceived health or lifestyle changes.
- Sociodemographic variables: age, gender, marital status, education, household income, ethnicity.
- Health-related variables: self-rated health, presence of chronic conditions, perceived weight status, frequency of healthcare visits.
- Behavioral and attitudinal variables (used as indicators of behavioral engagement): exercise frequency, enjoyment of exercise, willingness to share wearable data with a healthcare provider, intent to use a healthcare provider’s app on a smartwatch. In this study, behavioral engagement was defined as health-related behaviors and openness to using digital health tools.

The conceptual framework guiding the selection and grouping of variables is presented in Figure 1.

### Sample size considerations

A maximum of 3,000 participants was targeted. Assuming a prevalence of wearable device use between 25–30%, this sample size provided 95% confidence to estimate prevalence within ±3% precision. It also ensured adequate power to explore associations between sociodemographic, health, and behavioral variables and wearable use.

### Statistical analysis

Sociodemographic characteristics, health status and conditions, and behavioral factors of participants were described using frequency distributions. The prevalence of wearable health care device use was estimated, and associations with selected characteristics were initially evaluated using chi-square tests and univariable logistic regression models. Variables with a p-value ≤0.20 in univariable analyses were subsequently entered into a multivariable logistic regression model. Statistical significance in the multivariable model was determined at a p-value <0.05. Effect estimates were reported as odds ratios (ORs) and adjusted odds ratios (aORs), with corresponding 95% confidence intervals (CIs).

Multicollinearity was assessed using the variance inflation factor (VIF), with values ≥5 indicating multicollinearity (42). Across the selected characteristics, indeterminate responses (e.g. missing or blank entries) were excluded. As missing data were below 5%, an acceptable threshold for complete case analysis, (43, 44) no imputation was performed. Interactions were not investigated.

Descriptive and regression analyses were performed using Stata/SE version 18.0 (Stata Corporation, College Station, TX, USA).(45)

### Ethics

The study protocol was approved by the Weill Cornell Medicine in Qatar Institutional Review Board (WCM-Q IRB). The research was conducted in accordance with the principles of the declaration of Helsinki. Participation was voluntary, and all participants provided electronic informed consent prior to survey completion.

## Acknowledgements

This study was funded by Qatar Research, Development, and Innovation Council (QRDI) through the Undergraduate Research Experience Program (UREP) (Grant No. UREP30-102-3-033) The funder played no role in study design, data collection, analysis and interpretation of data, or the writing of this manuscript. The content of this publication is solely the responsibility of the authors and does not necessarily represent the official views of QRDI.

## Author contributions

HZ, BA, MA, AA, RA, OA, FA, MRMA, ZT, SZ, and TA contributed according to the Contributor Roles Taxonomy (CRediT) as follows:

**Conceptualization:** HZ, TA

**Methodology:** HZ, TA

**Investigation:** BA, MA, AA, RA, OA, FA, MRMA

**Data Curation:** HZ, ZT, SZ

**Formal Analysis:** HZ, ZT, SZ

**Writing - Original Draft:** HZ, BA, MA, AA, RA, OA, FA, MRMR, ZT, SZ, TA

**Writing - Review & Editing:** HZ, BA, MA, AA, RA, OA, FA, MRMA, ZT, SZ, TA

**Project Administration:** HZ, TA

**Supervision:** HZ, TA

**Funding Acquisition:** HZ, TA

## Data Availability

The datasets generated and/or analysed during the current study are not publicly available but are available from the corresponding author on reasonable request.

